# 3D Printed frames to enable reuse and improve the fit of N95 and KN95 respirators

**DOI:** 10.1101/2020.07.20.20151019

**Authors:** Malia McAvoy, Ai-Tram N. Bui, Christopher Hansen, Deborah Plana, Jordan T. Said, Zizi Yu, Helen Yang, Jacob Freake, Christopher Van, David Krikorian, Avilash Cramer, Leanne Smith, Liwei Jiang, Karen J. Lee, Sara J. Li, Brandon Beller, Michael Short, Sherry H. Yu, Arash Mostaghimi, Peter K. Sorger, Nicole R. LeBoeuf

**Affiliations:** Greater Boston Pandemic Fabrication Team (PanFab) c/o Harvard-MIT Center for Regulatory Science, Harvard Medical School, Boston, MA, USA; Harvard-MIT Division of Health Sciences and Technology, Cambridge, MA, USA; Harvard Medical School, Boston, MA, USA; Harvard Graduate School of Design, Cambridge, MA, USA; Harvard Ludwig Cancer Research Center and Department of Systems Biology, Harvard Medical School, Boston, MA, USA; Harvard-MIT Center for Regulatory Science, Harvard Medical School, Boston, MA, USA; Fikst Product Development, Woburn, MA, USA; Borobot, Middleborough, MA, USA; Dana-Farber Cancer Institute, Boston, MA; Department of Radiology, Brigham and Women’s Hospital, Boston, MA, USA; Department of Dermatology, Brigham and Women’s Hospital, Boston, MA, USA; Engineering Science at Norwalk Community College Norwalk, CT, USA; Department of Nuclear Science and Engineering, Massachusetts Institute of Technology, Cambridge, MA, USA; Department of Dermatology, Yale University School of Medicine, New Haven, CT USA; Harvard Program in Therapeutic Science, Harvard Medical School, Boston, MA, USA

**Keywords:** COVID-19, pandemic response, personal protective equipment (PPE), N95 respirators, KN95 masks, 3D printing, filtering face piece (FFP) respirator, mask frames, prototyping, occupational health

## Abstract

**Background:** In response to supply shortages during the COVID-19 pandemic, N95 filtering facepiece respirators (FFRs or “masks”), which are typically single-use devices in healthcare settings, are routinely being used for prolonged periods and in some cases decontaminated under “reuse” and “extended use” policies. However, the reusability of N95 masks is often limited by degradation or breakage of elastic head bands and issues with mask fit after repeated use. The purpose of this study was to develop a frame for N95 masks, using readily available materials and 3D printing, which could replace defective or broken bands and improve fit.

**Results:** An iterative design process yielded a mask frame consisting of two 3D-printed side pieces, malleable wire links that users press against their face, and cut lengths of elastic material that go around the head to hold the frame and mask in place. Volunteers (n= 41; average BMI= 25.5), of whom 31 were women, underwent qualitative fit with and without mask frames and one or more of four different brands of FFRs conforming to US N95 or Chinese KN95 standards. Masks passed qualitative fit testing in the absence of a frame at rates varying from 48 – 92% (depending on mask model and tester). For individuals for whom a mask passed testing, 75-100% (average = 86%) also passed testing with a frame holding the mask in place. Among users for whom a mask failed in initial fit testing, 41% passed using a frame. Success varied with mask model and across individuals.

**Conclusions:** The use of mask frames can prolong the lifespan of N95 and KN95 masks by serving as a substitute for broken or defective bands without adversely affecting fit. Frames also have the potential to improve fit for some individuals who cannot fit existing masks. Frames therefore represent a simple and inexpensive way of extending the life and utility of PPE in short supply. For clinicians and institutions interested in mask frames, designs and specifications are provided without restriction for use or modification. To ensure adequate performance in clinical settings, qualitative fit testing with user-specific masks and frames is required.

## BACKGROUND

Frontline health care workers are vulnerable to infection by the severe acute respiratory syndrome coronavirus 2 (SARS-CoV-2), which causes coronavirus disease 2019 (COVID-19)(1): among healthcare workers at the height of the pandemic in Wuhan, China, approximately 29% are thought to have acquired COVID-19 through hospital associated transmission (2). The most common routes of transmission for SARS-CoV-2 include: droplet transmission via direct face-to-face contact between patients and healthcare providers through coughing, sneezing, or speaking that aerosolizes virus, and aerosol-generating procedures such as intubation; indirect transmission by touching surfaces contaminated with virus followed by touching the face, nose or eyes is another suspected transmission route (3,4). Respiratory protection is an essential component of preventing hospital-based infections during the current COVID-19 pandemic, but an unprecedented demand for N95 filtering face mask respirators (FFRs; N95 masks) has led to severe shortages and many institutions have been forced to look for ways to prolong mask usability an even alternative solutions to respiratory protection.

The US Centers for Disease Control (CDC) recommends that healthcare workers dispose of N95 masks after each patient encounter. However, during the 2009 influenza A (H1N1) pandemic, which involved many fewer cases and deaths than the current COVID-19 pandemic, N95 respirator supplies were depleted (5,6). An evaluation of respiratory programs in California hospitals revealed that half of hospital managers interviewed (n=48) reported shortages of N95 respirators due to increased demand and slow delivery from suppliers (7). In response, guidelines were developed to preserve N95 supplies during shortages; these included “extended use” and “reuse.” “Extended use” is defined as the practice of wearing the same respirator for contact with several different patients infected with the same respiratory pathogen, without disposing of the respirator between patients. “Reuse” refers to the practice of using the same N95 respirator after removing it (“doffing”), for instance after a healthcare worker’s shift has ended, and then putting it back on (“donning”) prior to the next patient encounter. In the current COVID-19 pandemic, in which the shortage of PPE is more severe and widespread than in 2009, there is no specific regulation on the number donning/doffing cycles for N95 masks (8), although previous work found that masks consistently failed a test for fit quality after five consecutive donnings (9). Individual healthcare settings have therefore enacted their own policies, often with insufficient information, to restrict respirator reuse and extended use (10).

In the US, N95 FFRs are regulated by the National Institute for Occupational Safety and Health (NIOSH; part of the CDC) and the Food and Drug Administration (FDA) and must conform to standards set out in US 42 CFR part 84. Other countries have analogous regulations, including the GB2626-2006 standard for KN95 masks in China and the EN 149:2001 standard for FFP2 masks in Europe. All such masks must exhibit three essential properties: (i) efficient filtration of small particles (ii) unencumbered inhalation and exhalation with a mask in place and (iii) snug fit to the face of a user so that all inhaled air passes through the filtering fabric. Mask reuse is often limited by problems achieving good fit as a result of breakage or degradation of elastic bands that hold the mask in place. In addition, increasingly poor respirator fit with reuse of N95 masks occurs due to degradation in nose clips and other components required to seal a mask tightly to a user’s face (11).

The aim of this study was to develop a device to replace defective bands on N95 respirators without interfering with successful fit, thereby prolonging the lifespan of N95 respirators for both extended use and reuse. We also aimed to improve fit for individuals who failed baseline testing, thereby increasing the number of individuals who could benefit from low-cost respiratory protection. In the latter case, a specific aim was to improve the fit of KN95 masks, which are similar in performance to N95 FFRs (12) but are often observed to fail fit testing (13). We sought to use readily available materials and common 3D printing technology in a design that was made freely available for use or further modification.

## RESULTS

### Frame design and fabrication

Development of a modular mask frame model was inspired by the work of Dr. Christopher Wiles at the University of Connecticut, who has used 3D printed frames to enable use of alternative filter fabrics such as Halyard H600 sterilization wrap (14). We attempted to use the same relatively rigid 3D printed design to hold in place a standard 3M Model 8210 (St. Paul, MN USA) N95 industrial respirator. However, we found that the frame did not fit many individuals, particularly females with narrow faces. Previous research has shown that there are key facial dimensions affecting respirator fit (15). We therefore sought to develop a frame with flexible components that could be molded by a user to assist in optimizing fit along these dimensions. This was achieved by using two malleable wire components (made of copper, steel or aluminum) to link two rigid lateral PLA frames fabricated by 3D printing (**Figure 1**). The final design was the result of an iterative process, which consisted of multiple rounds of mask and prototype fit testing on volunteers (students and healthcare professionals) with design modifications made based on user feedback. Direct interaction between users and designers facilitated the process. Key features added in the iterative design process included the production of two frame sizes to improve fit to faces of difference sizes and shapes, the addition of clips to help secure the mask frame to the underlying respirator and decrease the likelihood of the frame falling off during use, and modifications to the location and length of the frame head bands to make donning and doffing easier. We freely provide all designs in standard electronic formats for use by others or for further modification.

**Figure 1.**
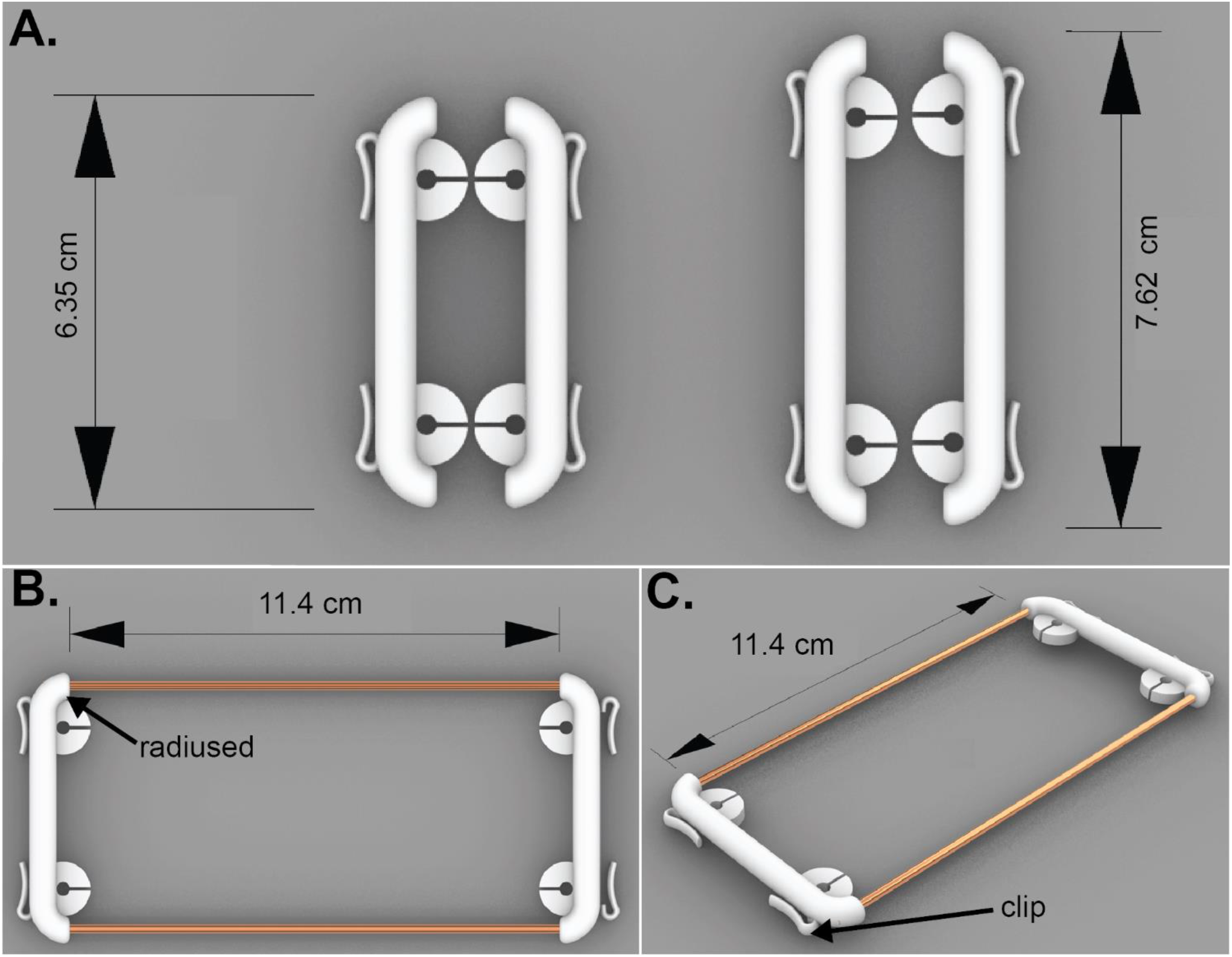
Mask frame components. **A)** PLA lateral frames in two sizes: the small size is 6.35 cm long and regular size is 7.62 cm long. **B and C)** Assembled mask frames consisting of both mask frame and malleable wire (copper, steel, or aluminum). Note that this mask frame involves attaching 3D printed components to wire using cyanoacrylate “superglue”. A mechanical attachment method is described in Figure 2.

A total of six weeks was required to design, prototype, and fabricate mask frames for testing. The resulting design was flexible enough to conform to a diversity of face types and sizes and also rigid enough to seal masks to users’ faces. Frames were fabricated from polylactic acid (PLA) on a standard 3D printer in less than 30 min at a cost of approximately 0.50 USD. Two sizes of lateral frames were printed in PLA and made available to participants: a “small” size (6.35 cm long) and a “regular” size (7.62 cm long) (**Figure 1A**). Additional features included radiused edges on the lateral frame **(Figure 1B)** to prevent the N95 FFR from being excessively deformed; deformation was observed with prototypes in which the edges were square or “V” shaped. We tested two different methods of attaching the malleable wire, one that used adhesive (**Figure 2A)** and one that involved twisting (**Figure 2B**). Note that slight modifications were necessary to accommodate 3D printed frames to the two wire attachment methods; however, the methods of wire attachment did not detectably affect comfort or function; design files for both prototypes are provided **(Additional Materials 1, 2, and 3)**.

**Figure 2.**
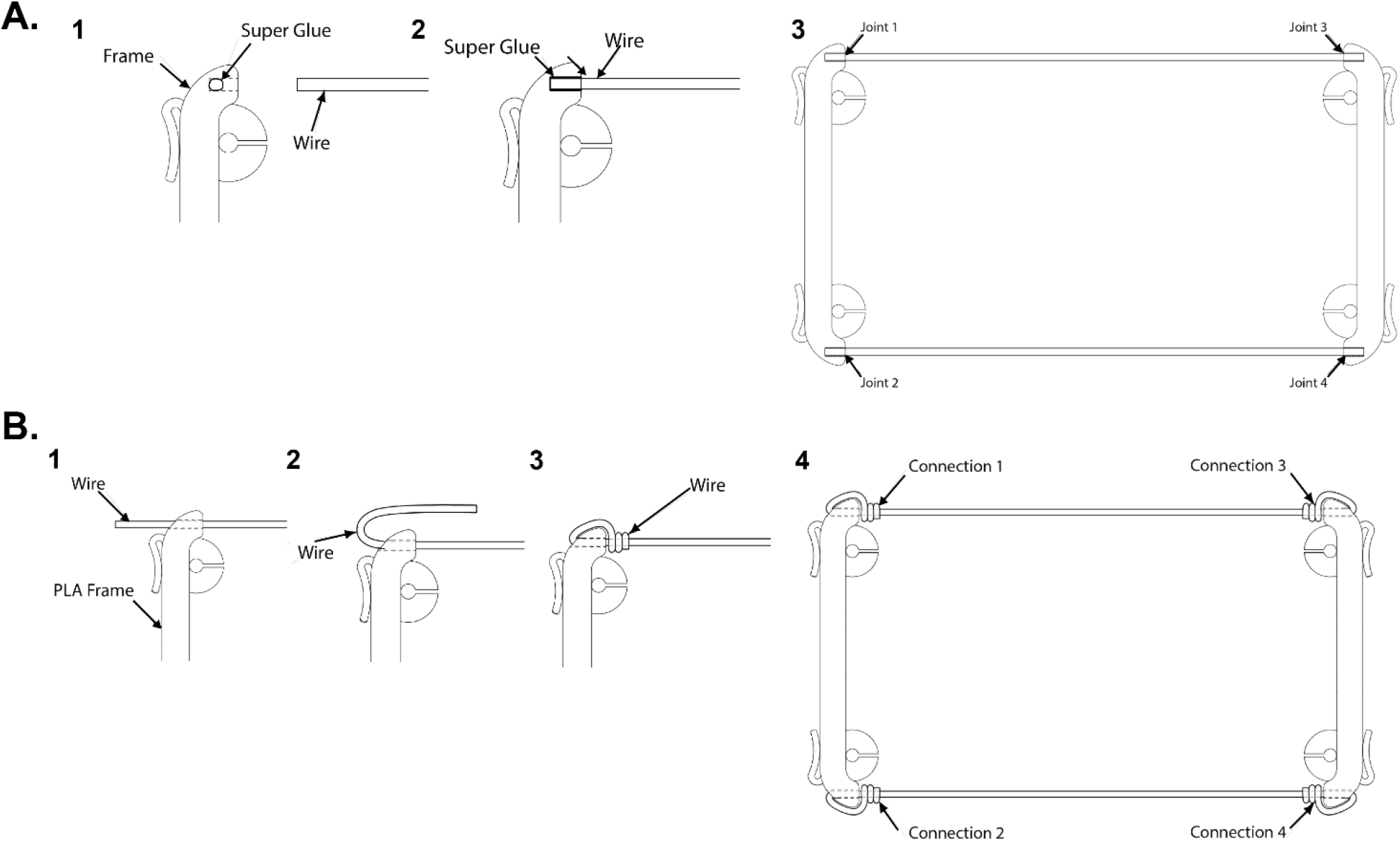
Methods for mask frame assembly. **A)** Method One for assembly of mask frame for N95 respirators utilizing glue adhesive. 1) One drop of cyanoacrylate superglue is placed into the end slot for the wire within the PLA lateral frame. 2) The end of a wire is inserted into the slot. 3) All four wire ends are inserted into the PLA slots as shown to complete the frame. **B)** Method Two for assembly of mask frame for N95 respirators using wire alone (no adhesive). 1) Wire is pushed through the opening in the PLA lateral frame. 2) The wire is looped back and 3) twisted around itself using pliers. 4) This process is repeated for each of the 4 total connections.

As a source of replacement elastic straps, we used non-latex phlebotomy tourniquets that are widely available and made of an FDA-approved material (Monprene^(R)^ PR-23040). Elastic straps lock into slots on each side of the 3D-printed piece and are cut to a standardized length that fits around users’ heads, just like factory- supplied straps (**Figure 3A, 3B**). Additional clips along the frame make it possible to secure any remaining respirator bands to the frame. These clips are designed to optimize the pressure holding the mask firmly in place (**Figure 1C, 3C**). A frame with an attached mask is donned just like a mask without a frame: the respirator is brought to the face to cover the nose and mouth, the lower strap is brought up and over the top of the head and the upper strap is pulled up behind the head. The nosepiece of the mask frame is then press-fit, or otherwise bent to the shape of a user’s nose (additional fitting instructions are provided in Methods; example fits shown in **Figure 4**). Individual users were allowed to choose a frame size based on whether their faces were “small” or “regular.”

**Figure 3.**
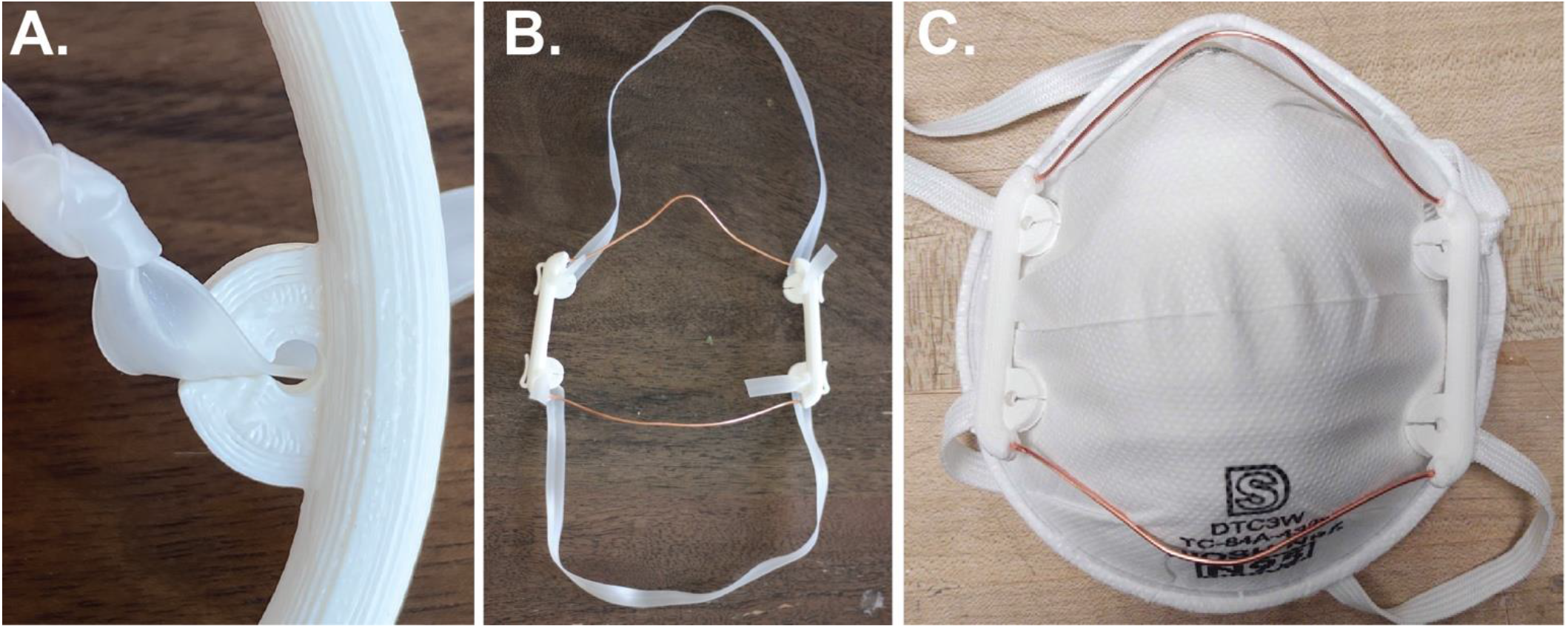
Attachment of the head band to a mask frame. **A)** A knot is tied at the end of each band and the band is then slid into and locked in place using the PLA slot **B)** Mask frame with bands **C)** Clip attachment of the frame to the bands on the 3M N95 Model 1860 respirator.

**Figure 4.**
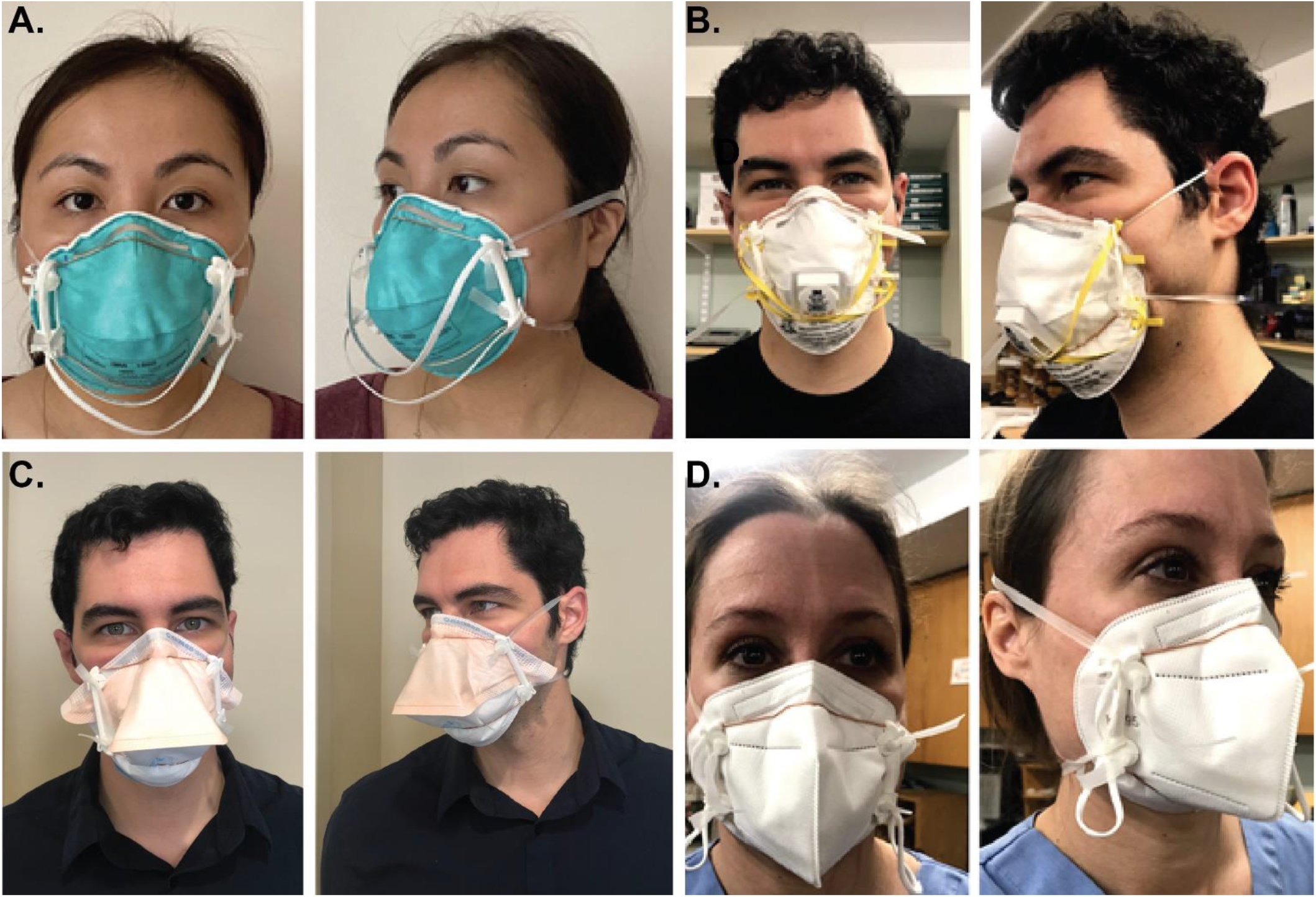
Properly donned mask frames and respirators on three different volunteers. **A)** A 3M 1860 N95 domed healthcare respirator, **B)** a 3M 8210 N95 domed industrial respirator (note the a valve-less version of the model is used in healthcare settings but was not always available for testing due to widespread respirator shortages) **C)** a Kimberly Clark duckbill, and **D)** a KN95 flat-fold respirator. The bands should be sufficiently tight and the nosepiece manipulated to achieve a good seal.

### Selection of FFRs for testing

Four different types of N95-style FFRs were selected for testing: a 3M Model 1860 N95, 3M Model 8210 N95, Kimberly Clark 46827/46767 (hereafter referred to as “duckbill”), and a Cooper KN95 (imprinted with number XK02-001-00010; Cooper USA; Los Angeles, CA). The 3M model 1860, available in both small and regular sizes, is a standard dome or cup-shaped respirator commonly used in healthcare settings (16) and is fabricated from media (fabric) that provides enhanced fluid and splash resistance (to ASTM Test Method F1862 (17)). The Kimberly Clark regular (46767) and small (46827) models, like the 3M 1860 model, were used in healthcare settings prior to the pandemic and are fabricated according to ASTM standards and but are duckbill shaped instead of dome-shaped. We also tested an industrial 3M model 8210 mask, only available in a single standard size, that would not normally be used in a healthcare setting but whose temporary use is permitted in the US under an FDA Emergency Use Authorization (EUA) issued on April 3, 2020 (18). The Cooper flat-fold KN95 respirator, available in one standard size, was selected as prototypical of a non-US manufactured FFRs whose use in healthcare is also allowed by an FDA EUA. Flat-fold respirators have a very different shape and fit from cup- style respirators. With KN95 masks it has also been observed that even when filtration efficiency meets specification fit can be problematic (19,20).

### Test subject demographics

A total of 41 volunteers were involved in this study and consisted of attending physicians, resident physicians, medical students, nurses, medical assistants, clinic staff, and research scientists, with predominantly female participants (ranging from 50% to 92% depending on the test group; **Table 1**). The proportion of female participants is representative of the healthcare workforce, which is predominantly female in the U.S. and worldwide (21). Of note, prior literature shows that women fail fit testing approximately 10% more frequently than men (22), suggesting a greater potential need for methods to improve fit with female FFR users. Individuals had a BMI ranging from 18.5 to 56.6 (averaging 25.5 for all groups). The study was approved by the Partners Healthcare Institutional Review Board (protocol 2020P001209) and all volunteers provided informed consent.

**Table 1.** Demographics and characteristics of participants undergoing baseline fit testing. SD = standard deviation.

| No. (%) or Mean +/- SD (range) |  |  |  |  |
| --- | --- | --- | --- | --- |
| Characteristic | 1860 N95 respirators<br>(n=32) | 8210 N95 respirators<br>(n=10) | KN95 Respirators<br>(n=25) | Kimberly Clark<br>duckbill<br>respirators<br>(n=13) |
| **Gender** |  |  |  |  |
| Female | 18 (56.3%) | 5 (50%) | 22 (88%) | 12<br>(92.3%) |
| Male | 14 (43.8%) | 5 (50%) | 3 (12%) | 1 (8.3%) |
| **Age** | 37.6 +/- 10.7 | 30.7 +/- 5.7 | 36.5 +/- 12.5 | 33.6 +/-<br>10.2 |
| **Ethnicity** |  |  |  |  |
| Asian/Pacific<br>Islander | 10 (31.3%) | 4 (40%) | 5 (20%) | 0 |
| White/Caucasian | 10 (31.3%) | 6 (60%) | 8 (32%) | 3 (25%) |
| Black/African<br>American | 10 (31.3%) | 0 (0%) | 9 (36%) | 8<br>(66.7%) |
| Hispanic/Latino | 2 (6.3%) | 0 | 3 (12%) | 2<br>(16.7%) |
| **Body mass index**<br>**(BMI)** | 25.7 +/- 7.8 | 23.4 +/-2.7 | 26.6 +/- 8.5 | 30.5 +/-<br>10.2 |
| **Healthcare role** |  |  |  |  |
| Attending physician | 16 (50%) | 1 (10%) | 6 (24%) | 1 (8.3%) |
| Resident physician | 0 | 2 (20%) | 2 (8%) | 0 |
| Medical student | 2 (6.3%) | 3 (30%) | 2 (8%) | 0 |
| Graduate student | 0 | 1 (10%) | 0 | 0 |
| Researcher | 2 (6.5%) | 3 (30%) | 1 (4%) | 0 |
| Nurse | 4 (12.5%) | 0 | 5 (20%) | 3 (25%) |
| Medical Assistant | 5 (15.6%) | 0 | 5 (20%) | 5<br>(38.5%) |
| Clinic Staff | 3 (9.4%) | 0 | 4 (16%) | 4 (30.8%) |
| **Mask size (previously fitted)** |  |  |  |  |
| Small | 10 (31.3%) | 5 (50%) | 6 (24%) | 1 (8.3%) |
| Regular | 22 (68.8%) | 5 (50%) | 19 (76%) | 12 (92.3%) |

### Qualitative FFR fit testing

Among the group of 41 volunteers, 32 were fit-tested with 3M model 1860 masks, 10 with 3M 8210, 25 with Cooper KN95 masks, and 13 with the Kimberly Clark duckbills; not all masks could be tested on all individuals due to severely limited mask supply resulting from the COVID-19 pandemic. When available, information provided by the participants about which mask size they had previously used in a clinical setting was used to guide the selection of an appropriate respirator and mask frame size for this study (in all cases previous experience was with a small or regular 3M model 1860; **Table 1)**. Qualitative fit testing was performed using a 3M FT14 standardized hood and 3M FT-32 bitter testing solution; if a user could taste the aerosolized fluid, the test was judged to have failed. Fit was tested without a mask frame (the baseline condition) and with the 3D printed mask frame in place of the mask straps (the test condition; outlined in **Figure 5)**. For 3M 8210 masks and Kimberly Clark duckbills, 9/10 (90%) and 12/13 (92.3%) of participants passed baseline testing without a 3D printed mask frame, respectively. For the 1860 and KN95 masks, baseline passing rates were lower at 28/32 (87.5%) and 12/25 (48.0%), respectively. The passing rates for 1860, duckbill, and 8210 masks are consistent with previous literature demonstrating 82-95% fit rates across N95 respirator models (22,23). We then asked whether individuals for whom a mask passed qualitative fit testing in the baseline condition would also pass fit testing when elastic straps were removed (or allowed to hang down) and the masks held in place with a frame and replacement straps. For the 8210 (**Table 2; Figure 4B**), 100% of participants who passed qualitative fit testing at baseline preserved fit using a mask frame (9/9). For individuals who passed fit testing with an 1860 mask (**Table 2; Figure 4A**), a KN95 mask (**Table 2; Figure 4D**), or a Kimberly Clark duckbill (**Table 2; Figure 4C**), the passing rates with the 3D mask frame in the absence of the original mask straps were 22/28 (78.6%), 11/12 (91.7%), and 9/12 (75.0%), respectively.

**Table 2.** Qualitative fit testing results using mask frames.

| Mask Type | Number Passed Qualitative Fit Test |  |  |
| --- | --- | --- | --- |
|  | Baseline Testing of Mask without Frame | Participants Who Passed Baseline: Mask with Frame (Preserving Fit) | Participants who Failed Baseline: Mask with Frame (Improving Inadequate Fit) |
| 8210 model (Total n=10) | 9/10 (90%) | 9/9 (100%) | 1/1 (100%) |
| 1860 model (Total n=32) | 28/32 (87.5%) | 22/28 (78.6%) | 0/4 (0%) |
| KN95 model (Total n=25) | 12/25 (48.0%) | 11/12 (91.7%) | 6/13 (46.2%) |
| Kimberly Clark duckbill model (Total n=13) | 12/13 (92.3%) | 9/12 (75.0%) | 0/1 (0%) |

**Figure 5.**
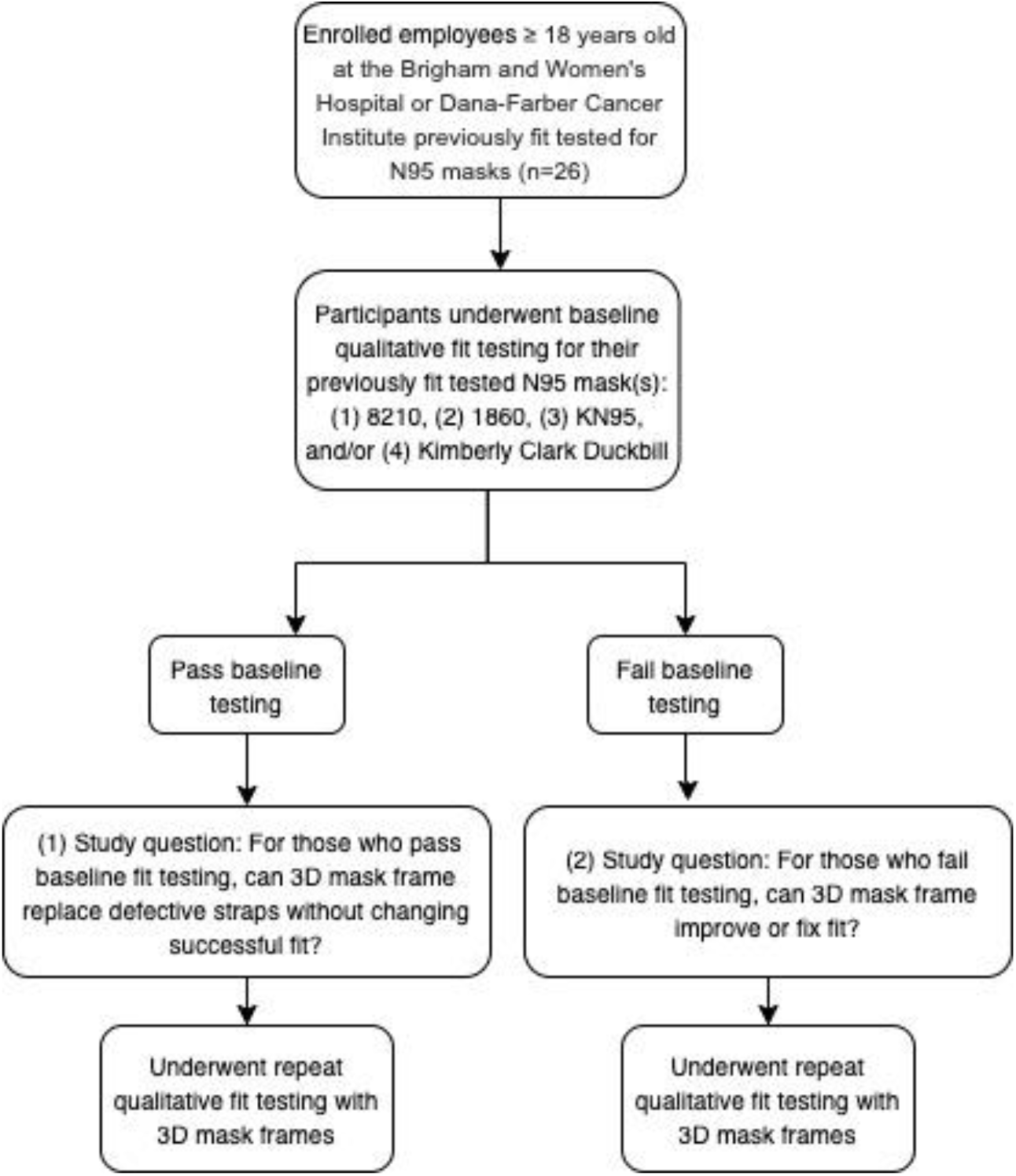
Flow chart of study methods.

We also asked whether mask frames could improve fit for participants who failed baseline testing. For the Model 8210 mask, use of a frame made it possible for a single participant who did not fit a mask at baseline to pass. However, in no case was a frame able to improve fit of an 1860 mask (0/4 participants) or a Kimberly Clark duckbills (0/1 participants). In contrast, 46.2% (6/13 participants) for whom the KN95 mask did not pass fit testing under baseline conditions achieved an acceptable fit with a frame (**Table 2**).

## DISCUSSION

In this paper we describe an iterative design process, involving multiple rounds of prototyping, clinical feedback, and design modification that resulted in a simple frame consisting of two identical 3D printed components (made in two sizes to accommodate different faces) and two pieces of malleable wire that together can hold an N95 or KN95 mask to users’ faces in the absence of factory-supplied straps. Such mask frames are reusable and can be sterilized using 70% isopropyl alcohol wipes. Across a diverse group of volunteers, we found that mask frames were effective in replacing the original straps on all three masks tested. Results were mixed with respect to our additional goal of improving fit for participants who failed baseline qualitative fit-testing: frames were effective for some individuals and mask models and ineffective in other cases. The most promising results were obtained with KN95 flat-fold masks, for which achieving a good fit is known to be challenging(13). Replacing elastic straps broken or degraded with age, multiple donning/doffing cycles, or repeated decontamination and increasing the number of individuals who can fit KN95 masks would immediately impact PPE supplies for healthcare workers.

Multiple recent projects have attempted to develop reusable respirators to replace disposable N95s, especially within the global 3D printing community. For instance, the Copper3D NanoHack mask is printed with a PLA filament as a flat piece, and is manually assembled into a 3D configuration using hot air (e.g. hairdryer) or hot water (24); two reusable filter cartridges are then inserted into an intake port. The HEPA Mask (25), Creality Mask (26) and the Lowell Makes Mask (27) all involve similar 3D printed components in PLA but with different variations in the filter holders. However, producing FFRs *de novo* involves several challenges, including securing a supply of suitable filters and achieving the proper fit for a wide range of users. This issue of fit has been tackled by printing masks in several sizes, experimenting with flexible materials or surface scanning an intended user’s face and creating a custom-fit device (World Advanced Saving Project (WASP)(28) and Bellus3D (29)). However, throughput is currently limited (30) and in some cases supplies of the necessary filters have been largely depleted (31). Thus, conventional N95 masks are likely to remain an essential component of PPE during the current pandemic (32).

How great is the need for extending usable mask life? Historical guidance by the National Institute for Occupational Safety (NIOSH) specifies that the useful lifetime of NIOSH-approved FFRs is limited by filter load and that any filter or mask should be replaced if it becomes soiled, damaged, or causes noticeably increased breathing resistance (33). In environments that generate high cumulative filter loading, the recommended maximum lifespan for N95 respirators is eight hours and it is standard practice in healthcare to dispose of N95 masks after each patient encounter. However, during the first SARS pandemic, the CDC stated that “health care facilities may consider reuse as long as the device has not been obviously soiled or damaged (e.g. creased or torn)” and “if a sufficient supply of respirators is not available (34).” Multiple FDA Emergency Use Authorizations (EUAs) issued during the COVID-19 pandemic have further expanded on this concept and led to the use of a wide variety of decontamination methods including UV germicidal irradiation, vaporous and ionized hydrogen peroxide (VHP/iHP) and moist heat (35–42), all of which promise to enable N95 reuse (for instance, the FDA EUA for the Battelle decontamination system permits up to 20 vaporous hydrogen peroxide decontamination cycles per respirator(43)). However, mask reuse is frequently limited by breakage or degradation of the elastic bands that hold a mask in place and it has been reported that N95 masks stored in preparation for a pandemic also have a high rate of band failure (11).

Moreover, quantitative fit testing (e.g. using a PortaCount quantitative fit testing apparatus, TSI Inc., Shoreview, MN) has shown that multiple donnings and doffings degrade fit independent of band failure. Bergman et al. (9) found that, after five consecutive donnings, fit factors consistently dropped below 100, the cut-off between passing and failing the test. Vuma et al. (44) showed that, when 25 tests subjects performed consecutive N95 donning and doffing operations, fit factor differed significantly between the first and the sixth re-donnings. After the sixth donning, only ∼68% of study subjects achieved a passing fit. Degsys et al. (45) found that an increase in fit failures was associated with an increasing number of shifts, each of which was associated with a donning and doffing cycle (median 4 shifts) and increasing hours of use (median 14 hours). It is thought that failures of fit with prolonged use of N95 masks involves degradation of the malleable nose clips and other components that help seal a mask tightly to a user’s face. Additionally, mask fit is adversely affected by repeated cycles of decontamination across a variety of methods (including heat and ethanol) (46). If undetected, poor fit causes air to flow around the mask (11) potentially increasing disease transmission.

Problems with fit are not restricted to masks that are being reused: it has previously been reported that even with new masks, about 17% of users will fail fit testing with any specific model of N95 or N95-equivalent mask (23). The fit failure rate for KN95s has not been extensively quantified in the literature, but available studies suggest that fit failure is an issue with a majority of KN95 models (40). Improving fit by using a frame would therefore be helpful even in non-pandemic situations. The problem with failure to fit any mask is made worse in a pandemic by shortages in alternative forms of respiratory protection (e.g., powered air-purifying respirators(47)). Thus, both pandemic and non-pandemic respiratory protection presents a substantial need for devices – such as the mask frames described here - to extend the useful lifetime of FFRs, such as N95 or KN95 masks, or improve fit of new masks. It must be noted, however, that clinical data supporting extended use and reuse of N95 masks, with or without decontamination, remains limited. Concerns about extended use and reuse involve not only fit and the adequacy of the seal to a user’s face (45), but also potential infection risks acquired during donning/doffing (since the outer surface of respirators can be contaminated with infectious agents that can be transferred to a user (48)) and reductions in filtration efficiency over time. The mask frames described here address only the first of these issues.

### Limitations of this study

This study has several limitations; most notably, that sample sizes and the number of mask models tested were small. We were unable to follow up preliminary but promising data that frames can improve the fit of KN95 masks. These limitations reflect continuing shortage in FFRs of all types and our inability to divert more than a small number of masks from hospital or staff use to a research project. In particular, results would be improved by finding a much larger number of participants who failed baseline fit testing and from whom multiple models of masks could be evaluated with and without frames. For all of the masks used in this study straps were artificially broken or allowed to hang free; to better represent the real-world use case it will be necessary to perform fit testing with and without a mask frame after extended N95 mask use in a clinical setting. Our data suggest that the precise shape of a mask and the properties of the material may determine whether a frame can successfully substitute for original straps or improve fit. Additional research will be required to identify these variables and address them. Ideally, all of these issues will become increasingly possible to address as supply disruptions recede and sufficient FFRs can be dedicated to research studies.

## CONCLUSIONS

The use of the 3D-printed mask frames described in this study can prolong the lifespan of N95 and KN95 masks by serving as a substitute for broken or defective bands without adversely affecting fit. Frames also have the potential to improve fit for some individuals who cannot use existing masks. Both defective straps and poor fit are limiting factors in extended mask use and in the reuse of masks after decontamination, as masks with either defect are currently discarded. Thus, improving mask fit through use of a frame is expected to help offset urgent respirator supply shortages during the COVID-19 pandemic and also help after the pandemic passes. All design files and testing results are included in this manuscript and available for reuse without restriction; design information is also available via the PanFab website (https://www.panfab.org/). Our prototyping and testing efforts took place over the span of approximately 6 weeks and use of our designs should save substantial time relative to designing mask frames *de novo*. However we suggest that each group perform its own fit testing with a representative group of users.We believe that the results and designs presented here are one step in improving the supply and effectiveness of PPE in this pandemic and in increasing our collective ability to respond to future healthcare crises.

## METHODS

### Mask Frame Software and Design

3D printed mask frames were designed in Rhinoceros^(R)^ Rhino 6 (**Figure 1**) in two sizes. A 3D model (.3dm) of the lateral frames were exported in Rhino 6 to a Standard Tessellation Language (.STL) file. The .STL was uploaded to 3dPrinterOS, a cloud based 3D printing service. 3dPrinterOS converted the .STL to a G-code file, which contains machine commands that control the 3D printers’ movement and deposition of material, and sent to a 3D printer. Print settings were chosen by using the default values for the 3dPrinterOS customized for the Dremel 3d45 3D printer, including print nozzle temperature of 230°C and print bed temperature of 60°C. Other printer settings included a standard layer height of .3mm, a 1.2mm sidewall shell thickness, 10% infill in a ‘grid’ pattern, and a top and bottom layer shell thickness of 2mm.

### 3D Printer Model and Hardware

Dremel branded 1.75 mm diameter PLA was used on a Dremel 3d45 3D printer for printing all components. The printer has a .4mm nozzle extrusion width, and a build volume of 10 × 6.7 × 6 inches (254 × 152 × 170 mm). Print time for one regular sized mask frame was ∼ 30 min.

### Mask Frame Assembly

Two methods of mask frame assembly were used, each of which involved a slight modification to the 3D printed lateral frame. **Method 1** (**Figure 2A**) used an adhesive, cyanoacrylate (also known as super glue), to join the mask frame components; the prototype design is shown in **Figure 1**. Method 1 assembly sequence is as follows: gather components (2 flexible wires cut to 127 mm, 2 PLA lateral frames, and 1 bottle of superglue). (1) Place one drop of super glue in the lateral frame joint. (2) Insert wire into joint. Follow instructions accompanying super glue for holding wire in place to properly allow the glue to set and cure. (3) Repeat for each joint.

**Method 2** (**Figure 2B**) involved a mechanical connection in which formable wire was twisted to join mask frame components. Method 2 assembly sequence is as follows: gather components (2 flexible wires cut to 195 mm and two PLA lateral frames that have a hole through the joint). (1) Push wire through the joint in the lateral frame. (2) Loop wire back. (3) Twist wire around itself. Pliers are recommended to assist in bending and twisting of wire to ensure secure twist. (4) Repeat for each connection.

### Band and Clip Attachment

Based on prior work creating 3D printed mask frames by colleagues at the University of Connecticut (14), the band material used for this study was Monprene^(R)^ PR-23040 in the following size: 0.25 in × 0.015 in (Teknor Apex; Pawtucket, RI). Two strips of elastic 305 and 330 mm in length were cut for the 1860 and KN95 masks, and two strips 356 and 381 mm in length were cut for the 8210. A knot was tied at each end of each band, approximately 25 mm from the end of the band. The knot locks into each slot in the PLA frame as shown in **Figure 3A** and **Figure 3B**. The clips along the PLA frame secure to existing bands of the respirator, if still present, to secure the frame to the mask (**Figure 3C**).

### Donning, Doffing and Sterilization

Once the mask frame is attached to the respirator using the clips, the respirator is donned just like a respirator without the frame. Holding the respirator and mask frame in the palms of the hands, the respirator is brought to the face to cover the nose and mouth, and the mask frame should not pass outside the borders of the mask. The bottom strap attached to the mask frame is brought up and over the top of the head and placed at the nape of the neck below the ears. The upper strap is pulled up behind the head and placed at the crown of the head. Then, the nosepiece of the mask frame is manipulated in the shape of the user’s nose until a secure seal and good fit are achieved (**Figure 4**). A seal check is performed by placing both hands over the mask and exhaling. If air leakage is observed, there is not a proper seal. Re-adjusting the nosepiece or pulling the straps tighter should be attempted until a proper seal is obtained. Mask frames can be sterilized using 70% isopropyl alcohol wipes.

### Qualitative Fit Testing

This study was approved by the Partners Healthcare Institutional Review Board (protocol #2020P001209). All subjects underwent qualitative fit testing at the Dana-Farber Cancer Institute during May-June, 2020. Participants included attending physicians, medical students and researchers. Qualitative fit testing using a 3M FT14 hood and 3M FT-32 bitter testing solution was performed over two different testing sessions, both consisting of tests without the mask frame (baseline) and with the 3D printed mask frame (**Figure 5)**. Qualitative fit failure occurred if the participant could taste the solution (bitter taste). Four different models of respirators were tested: 1860, 8210, duckbill, and KN95. Data was analyzed using Prism version 8 (GraphPad).

## Data Availability

All data generated or analyzed during this study are included in this published article and its supplementary information files.

## ABBREVIATIONS

3D: 3 dimensional
FFRs: Filtering Facepiece Respirators
IRB: Institutional Review Board
PLA: Polylactic Acid
PPE: Personal Protective Equipment

## DECLARATIONS

### Ethics approval and consent to participate

The Partners Healthcare Institutional Review Board approved this study (protocol #2020P001209).

### Consent for publication

Consent for publication has been obtained from the study participants. Written informed consent for publication was obtained from the study participants pictured in **Figure 4**. A copy of the consent form is available for review by the Editor of this journal.

### Author Contributions

Mask frame design prototyping and manufacturing: M.M., C.H., T.B, D.P, J.F, C.V., A.C., L.S., L.J., B.B., S.Y.

Mask frame clinical testing: M.M., T.B., J.S., Z.Y., H.Y., D.K., K.L., S.J.L., M.S., S.Y., A.M.

Writing: M.M., C.H., T.B., D.P., P.K.S., N.L.

Greater Boston Pandemic Fabrication (PanFab) Consortium Coordination: D.P., H.Y., P.K.S., N.L.

## Acknowledgements

Above all we thank the members of the Greater Boston Pandemic Fabrication Team (PanFab) for technical, administrative, and logistical support necessary for the execution of this project. Membership found at https://www.panfab.org/the-team-and-the-project/consortium-members.

Additional support was given by Harvard University Graduate School of Design, where 3D printing for the project occurred, in addition to providing access to design software and materials for prototyping.

## ADDITIONAL MATERIALS

Additional Material 1: .STL file for mask frame design corresponding to adhesive attachment method, “PanFab- MaskFrame-RigidLateralFrame-AdhesiveConnection,stl”

Additional Material 2: .STLfile for mask frame design corresponding to mechanical attachment method, “PanFab- MaskFrame-RigidLateralFrame-MechanicalConnection.stl”

Additional Material 3: .3dm file for all components all the components of mask frame prototypes, annotated in 3D, “PanFab-MaskFrame-Assembled.3dm”

